# Clinical and haematological correlates of severe dengue using 2023 epidemic data: a multicentre tertiary hospital-based analysis from Dhaka, Bangladesh

**DOI:** 10.1101/2025.11.12.25339750

**Authors:** M. Pear Hossain, Rudbar Mahmood, Farzana Sultana Bari, Afrin Naher Kana, Bibek Ahamed, Munia Afroza Shanta, Sadia Afrin, Musleha Akter, Tahmina Akter Anika, Mehejabin Nurunnahar, Sayeda Jahin Tasnim, Abu Bakkar Siddique, Linyan Li, Sheikh Taslim Ali

**Affiliations:** WHO Collaborating Centre for Infectious Disease Epidemiology and Control, School of Public Health, Li Ka Shing Faculty of Medicine, The University of Hong Kong, Hong Kong Special Administrative Region, China; Laboratory of Data Discovery for Health Limited, Hong Kong Science Park, New Territories, Hong Kong; Department of Public health, North South University, Dhaka 1229, Bangladesh; Department of Food Engineering and Nutrition Science, State University of Bangladesh, Narayanganj, Dhaka-1461, Bangladesh; Department of Statistics, Jahangirnagar University, Savar, Dhaka, Bangladesh; Infectious Diseases Division, International Centre for Diarrhoeal Disease Research, Bangladesh, Mohakhali, Dhaka, Bangladesh; Maternal and Child Health Division, International Centre for Diarrhoeal Disease Research, Bangladesh, Mohakhali, Dhaka, Bangladesh; Institute of Epidemiology, Disease Control and Research, Dhaka, Bangladesh; Department of statistics, Eden Mohila College, Dhaka, Bangladesh; Department of Data Science, City University of Hong Kong, Hong Kong SAR, China; Department of Infectious Diseases and Public Health, Jockey Club College of Veterinary Medicine and Life Sciences, City University of Hong Kong, Hong Kong SAR, China

**Author notes:** Communicating author (STA), The University of Hong Kong, Hong Kong Special Administrative Region, China.

## Abstract

**Background:** Dengue fever, caused by the dengue virus (DENV), is a significant public health threat in tropical and subtropical regions. The 2023 outbreak in Dhaka City, Bangladesh, resulted in over 321,000 reported cases and 1,705 deaths, highlighting the urgent need for understanding factors associated with disease severity.

**Methods:** This multi-center, hospital-based study analyzed 466 laboratory-confirmed dengue cases admitted to four tertiary hospitals in Dhaka during the peak epidemic period (September 1 to October 31, 2023). Demographic, clinical, and laboratory data were collected using a semi-structured questionnaire. Cases were categorized based on severity, and statistical analyses were performed to identify predictors of severe dengue.

**Results:** Severe dengue was associated with older age (OR = 1.77; p <0.001), blood transfusion requirements (OR = 15.26; p <0.001), elevated hematocrit (OR = 15.16; p <0.001), and lower body mass index (BMI) (OR = 0.11; p <0.001). Delays in seeking medical care significantly correlated with increased severity (p = 0.007). Symptomatic markers such as abdominal pain (p < 0.001) and respiratory distress (p = 0.013) were strong indicators of severe outcomes.

**Conclusions:** Our findings underscore the importance of timely medical intervention and highlight demographic and clinical factors that may contribute to severe dengue progression. The association between lower BMI and disease severity warrants further investigation. These insights can inform targeted public health strategies and improve clinical management during dengue outbreaks.

**Author Summary:** Dengue poses a significant public health challenge in Bangladesh. To better understand the epidemiological and clinical characteristics of dengue in Dhaka city, we conducted a study on 466 patients admitted to four distinct tertiary hospitals during the 2023 outbreak. Dengue cases were identified through serological testing for the presence of the NS1 antigen. Our analysis revealed that older age, the need for blood transfusions, elevated hematocrit levels, and lower body mass index were significant predictors of severe dengue outcomes. Furthermore, delays in seeking medical care were strongly correlated with increased disease severity, highlighting the critical importance of timely intervention. Symptoms such as abdominal pain and respiratory distress emerged as strong indicators of severe dengue. These findings emphasize the complex interplay of various factors in determining dengue severity and underscore the need for context-specific public health strategies. By identifying these predictors, healthcare providers can enhance the management of severe dengue cases and reduce the impact of future outbreaks.

## Introduction

Dengue fever, a mosquito-borne viral disease caused by the dengue virus (DENV), remains a major global public health threat, particularly in tropical and subtropical regions [1]. With an estimated 390 million infections annually, dengue imposes a substantial burden on healthcare systems, especially in endemic areas such as South and Southeast Asia [2]. In Bangladesh, dengue has emerged as a major health concern, with recurrent outbreaks and increasing severity in recent years. In 2023, the dengue outbreak was particularly catastrophic, with over 321179 reported cases and 1,705 deaths, highlighting the urgent need for a deeper understanding of factors driving severe disease progression [3].

Severe dengue, as defined by the World Health Organization (WHO) 2009 guidelines, is characterized by plasma leakage, severe hemorrhage, and organ impairment, often leading to life-threatening complications such as shock and multi-organ failure [4]. While the majority of dengue cases are mild or asymptomatic, early identifying of severity predictors is vital for timely interventions and resource allocation. Established risk factors include older age, comorbidities (e.g., diabetes, mellitus and hypertension), and delays in seeking medical care [5-9]. However, the role of body mass index (BMI) in dengue severity remains controversial, with some studies suggesting obesity as a risk factor and others reporting no significant association [8, 10-12]. This discrepancy underscores the need for population-specific studies to clarify the relationship between metabolic health and dengue outcomes.

Dhaka City emerged as the epicentre of dengue outbreak in 2023, contributing 34% of national cases and 57% of deaths in Bangladesh, provided a unique opportunity to investigate various risk factors for disease severity [3]. Despite the high burden of dengue in this region, limited data exist on the demographic, clinical, and laboratory profiles associated with severe outcomes, particularly during epidemic periods. Understanding these factors is essential for improving clinical management, guiding public health interventions, and reducing mortality in resource-limited settings.

This study aimed to identify key predictors of severe dengue using data collected from patients upon hospital admission during the 2023 outbreak in Dhaka City. By analyzing demographic characteristics, clinical timelines, comorbidities, and laboratory parameters, we sought to provide insights into the factors driving disease severity and inform targeted strategies for patient care and outbreak response. Our findings contribute to the epidemiological evidence on dengue risk factors and highlight the importance of target-specific outbreak mitigation strategies to address context-specific dengue transmission dynamics.

## Methods

### Study design, setting and participants

This multi-canter study was designed as to retrieve the individual-level epidemiological data on dengue fever. It was conducted to get a broader view of perception on different demographic, clinical and laboratory parameters that might affect the health condition of the patients. The study focused a subset of patients admitted to dengue wards in four tertiary hospitals in Dhaka City, Bangladesh. These hospitals included three branches of Islami Bank Hospital located in Kakrail, Mugda, and Mirpur, as well as Enam Medical College in Savar. Data were collected from the patients during the peak time (between 1 September and 31 October 2023) of 2023 epidemic in Dhaka.

A total of 466 laboratory-confirmed dengue cases were retrospectively reviewed from the dengue wards of these four hospitals. Prior to conducting the study, written consent was obtained from each hospital’s authority to access the dengue wards. A quantitative cross-sectional descriptive study was then carried out on individuals diagnosed with dengue, specifically identified through serological testing for the presence of the NS1 antigen. Information was collected from the patients directly and from the medical records using a pre-designed semi-structured questionnaire. Additionally, written consent was obtained from each patient before conducting the interview.

### Defining dengue cases and other variables

Dengue cases were categorized as dengue (with or without warning signs) and severe dengue, following the WHO 2009 dengue classification scheme [4]. Dengue encompasses a spectrum of symptoms, including fever, headache, muscle and joint pain, rash, and mild bleeding, with some patients exhibiting warning signs that indicate a higher risk of severe disease. Severe dengue was identified by meeting at least one of the following criteria: (a) *Severe plasma leakage*, determined by significant fluid buildup in the body, which can lead to complications such as swelling in the abdomen or lungs. This condition may be accompanied by an increased heart rate, low blood pressure and/or a narrow difference between systolic and diastolic blood pressure, suggesting critical drop in blood volume. (b) *Severe hemorrhage*, involves serious bleeding that can pose life-threatening risk. Symptoms may include vomiting blood, passing fresh blood in the stool, or having dark, tarry stools. These bleeding episodes are often associated with unstable blood pressure and a rapid drop in hemoglobin levels, further complicating the patient’s condition. (c) *Severe organ impairment*, signifies the dysfunction of vital organs and can manifest as confusion or altered consciousness, severe liver damage (indicated by significantly elevated levels of liver enzymes, ALT or AST), or inflammation of the heart muscle (myocarditis) [13]. Body mass index (BMI) is classified into three categories: underweight (<18.5), normal (18.5-24.9), and overweight (≥25). The length of hospital stay is the period from admission to discharge. Patients with previous medical conditions such as COPD, Asthma, Hypertension, Diabetes mellitus, Heart disease, or other comorbidities are considered to have comorbidities.

### Statistical analysis

The included patients were categorized into two groups based on the case severity (i.e., severe and non-severe dengue cases), and suitable statistical tests were performed to compare. Categorical variables were compared using the Chi-squared test, while continuous variables were compared using the parametric t-test and non-parametric Manne-Whitney U test upon validation of normality assumptions. A 2-tailed p-value less than 0.05 was considered statistically significant.

In order to check the dynamics of severity profiles in different clinical phases (febrile, critical and recovery phases), clinical and laboratory variables were observed and compared overtime with reference to the symptom onset. This comparison was made between the severe and non-severe dengue groups using Manne-Whitney U test for each day and by group overall.

We further assessed predictors of disease severity using two modeling approaches. Generalized linear mixed-effects models with a binomial link function were applied to clinical and laboratory variables with repeated measurements during the stay in hospital. Random effects accounted for within-patient correlations and inter-hospital variability. Logistic regression was used for variables measured at the admission. Both models were adjusted for demographic characteristics including age, BMI and the need for blood or platelet transfusions during the stay in the hospital.

### Ethical declaration

The Institutional Ethical Approval Committee of Primeasia University, Bangladesh (PAU/IEAC/23/123) reviewed and approved the study. Prior to conducting the interviews, written consent was obtained from each patient.

## Results

In 2023, a severe dengue outbreak caused an alarming number of cases and fatalities, with 321179 cases and 1705 deaths reported (Fig 1A & B). The situation was particularly critical in Dhaka city, where 110008 cases (34% of the total reported cases in Bangladesh) and 980 deaths (57%) occurred (Fig 1C-E). This study analyzed data collected during the peak period of the outbreak (September 1 to October 31, 2023) and focused on the demographic, clinical, and laboratory information of 466 patients admitted to four hospitals in Dhaka city (*details in Methods section*). After cleaning the data to confirm NS1 or IgM positivity, we could include 449 patients for conducting the main analysis in the study.

**Figure 1.**
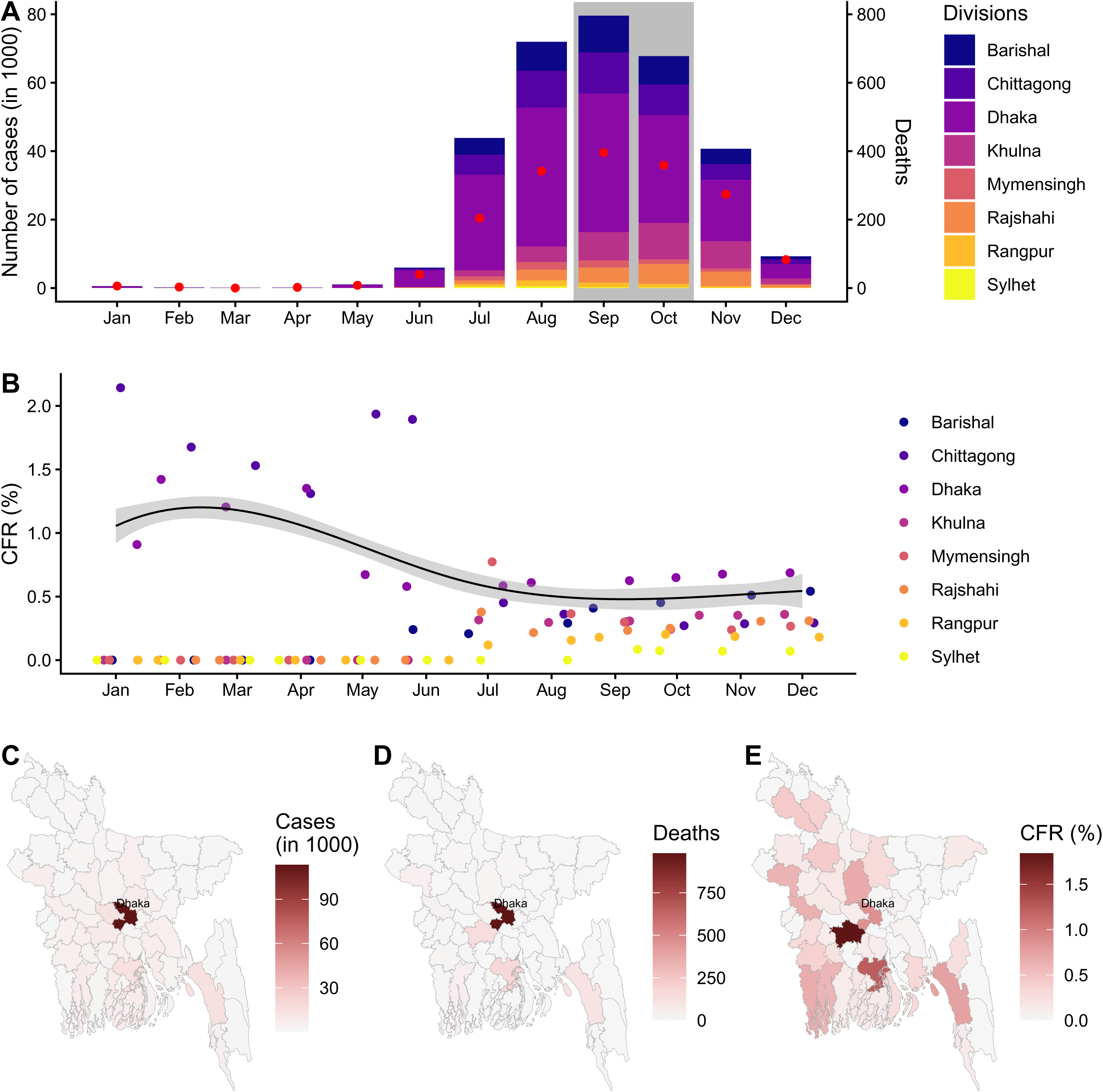
Dynamics of dengue outbreak in Bangladesh, 2023. (A) Monthly dengue cases across the eight primary administrative regions (divisions) of Bangladesh. The red dots in each month indicate the total number of deaths in Bangladesh for 2023. The grey shaded region indicates the period during which data were collected from individual patients upon hospital admission. (B) Case Fatality Rate (CFR, in %) by month and division, calculated as the percentage of cumulative deaths in each month divided by cumulative cases in that month. The solid line represents the overall CFR with 95% CI (grey shade) for Bangladesh across the months. (C) Spatial distribution of dengue cases across districts in Bangladesh, highlighting Dhaka district (second-level administrative region) as the area reporting the highest number of cases. (D) Spatial distribution of dengue-related deaths, with Dhaka district again showing the highest numbers. (E) Spatial distribution of CFR (%) across districts, with Faridpur district exhibiting the highest CFR, located adjacent to Dhaka district.

### Demographic and clinical characteristics

Cases of severe dengue found to be associated with a higher mean age compared to non-severe cases (39 ± 18 vs. 32 ± 18 years; *p* = 0.006), with a significant age-dependent trend in disease severity. The proportion of severe cases increased progressively across age groups, reaching 24% (9/38) among individuals aged >60 years, followed by 18% (17/93) in those aged 41–60 years, 15% (34/224) in the 19–40 years, and 6% (6/94) in individuals aged ≤18 years (*p* = 0.035) (Table S1). A significant higher prevalence were found for overweight individuals (BMI ≥25 kg/m²: 23% of severe cases vs. 13% in normal BMI and 3.2% in underweight; *p* = 0.022). Severe cases also exhibited longer mean onset-to-admission times (4.71 ± 1.88 days) than non-severe cases (3.99 ± 2.17 days; *p* = 0.003). Similarly, mean recovery times with reference to symptom onset were significantly longer for severe cases (8.72 ± 2.31 days) versus non-severe cases (7.68 ± 2.48 days; *p* < 0.001).

### Comorbidities and Disease Severity

The presence of any comorbidity significantly increased the likelihood of severe dengue outcomes (20% vs. 13% in non-severe cases; p = 0.039). While diabetes mellitus (22% vs. 14%; p = 0.10) and hypertension (20% vs. 14%; p = 0.2) demonstrated elevated severity rates, these associations were not statistically significant. No significant associations were observed for COPD/asthma (p = 0.502), heart disease (p = 0.548), or other comorbidities (p = 0.3) (Table S2).

### Symptomatic Markers of Severity

Abdominal pain (p < 0.001) and itchiness (p < 0.001) were strongly associated with severe outcomes. Respiratory distress (p = 0.013), joint pain (p = 0.026), and headache (p = 0.046) also showed significant associations with severe dengue. In contrast, fever (p = 0.5), rash (p = 0.4), and vomiting (p = 0.3) were found to be insignificant associations with severity. (Table S3).

### Temporal profile of clinical factors during time-since-symptom-onsets

Significant differences were observed in log-transformed platelet levels between severe and non-severe dengue cases with clear differences during 3-7 days of post-symptom onset (Fig 2A). The severe dengue cases exhibited lower platelet levels during this period. The lowest log-transformed platelet levels were observed around day 6 of post-symptom onset in both groups. Hematocrit levels also showed significant differences between severe and non-severe dengue cases (Fig 2B). Specifically, hematocrit percentages were lower in the severe dengue group between 9 and 11 days post-symptom onset. Both severity groups showed a progressive decline in hematocrit between days 5 and 7 post-onset. No statistically significant differences were observed in WBC count trajectories or SpO2 measurements between severity groups. The vital signs monitoring revealed significantly elevated SBP and DBP in severe dengue patients compared to non-severe cases within the first week, specifically on days 4–7 post-symptom onset (Fig 3A & B). However, SBP was lower among the severe group between 10 and 12 days post-symptom onset. Peak SBP occurred on day 7 in the severe group and on day 11 in the non-severe group. Liver function parameters showed no significant variation between severe and non-severe dengue cases (Fig 4).

**Figure 2.**
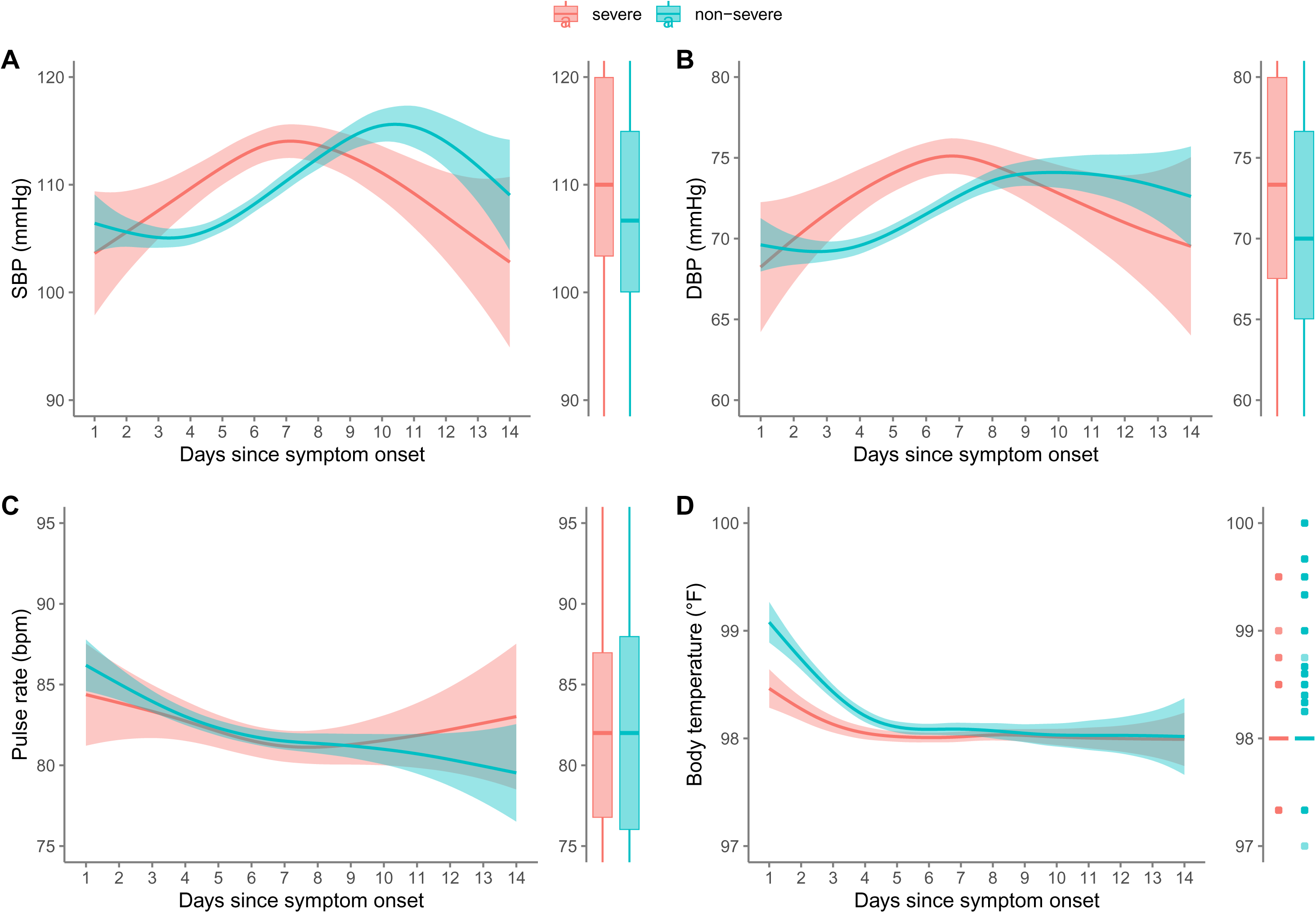
Dynamics of hematological profile among dengue patients admitted to the hospital. (A) Platelet counts measured in per microlitre blood (*μL*^-1^), (B) haematocrit measured in (%), (C) white blood cell count measured in per microlitre blood (*μL*^-1^), and (D) oxygen saturation (%). In each panel, lines and shaded regions indicate the fitted line obtained using generalized additive models, along with 95% confidence intervals (CIs). The boxplot in each panel represents the distributions of hematological profile categorized by severity levels. Asterisks denotes the significant p-values from the Mann-Whitney U test comparing the hematological profiles between two groups of dengue patients: severe vs non-severe.

**Figure 3.**
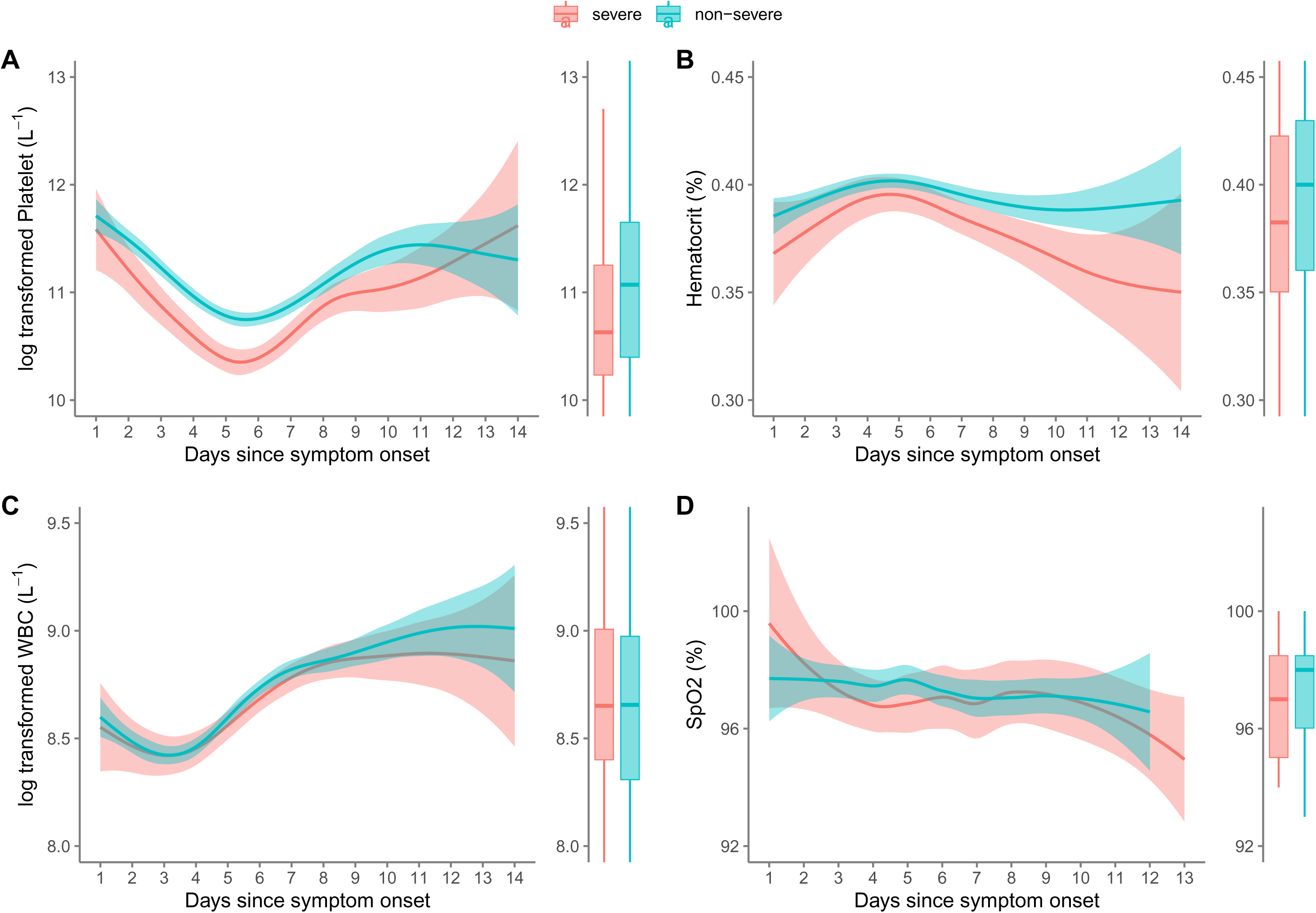
Dynamics of vital signs monitoring among dengue patients admitted to the hospital. (A) Systolic blood pressure measured in mmHg, (B) diastolic blood pressure measured in mmHg, (C) pulse rate measured in bpm (beats per minute), and (D) body temperature (℉). In each panel, lines and shaded regions indicate the fitted line obtained using generalized additive models, along with 95% confidence intervals (CIs). The boxplot in each panel represents the distributions of vital signs categorized by severity levels. Asterisks denotes the significant p-values from the Mann-Whitney U test comparing vital signs between two groups of dengue patients: severe vs non-severe.

**Figure 4.**
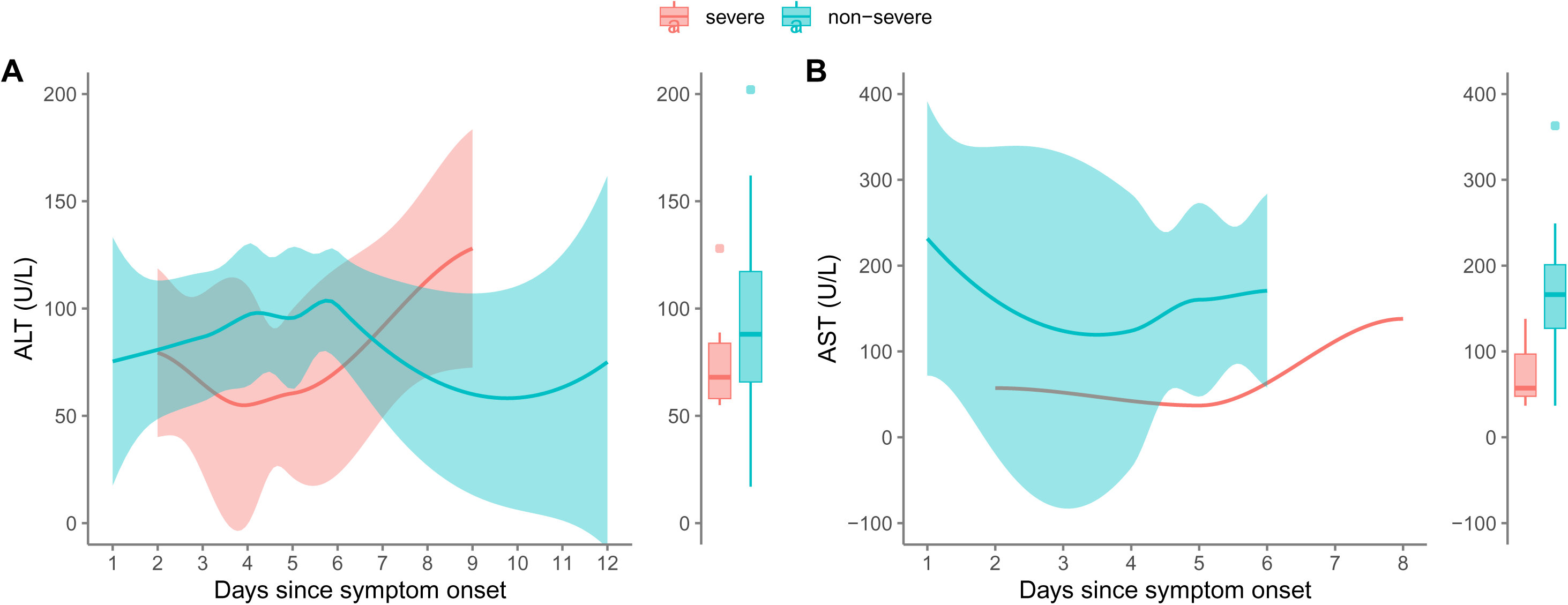
Dynamics of liver function tests among dengue patients admitted to the hospital. (A) ALT (alanine aminotransferase) and (B) AST (aspartate aminotransferase) measured in units per liter (U/L). In each panel, lines and shaded regions indicate the fitted line obtained using generalized additive models, along with 95% confidence intervals (CIs). The boxplot in each panel represents the distributions of liver function tests categorized by severity levels. Asterisks denotes the significant p-value from the Mann-Whitney U test comparing liver function tests over time between two groups of dengue patients: severe vs non-severe.

### Factors affecting severity of dengue

The generalized linear mixed-effects model identified significant predictors of dengue severity. The requirements for blood or platelet transfusions (OR = 15.26, p <0.001) and elevated haematocrits levels (OR = 15.16, p <0.001) were associated with the dengue severity, indicating that these factors increase the odds of severe dengue by more than fifteenfold (Table 1). Older age (OR = 1.77 per year, p <0.001) and diastolic blood pressure (OR = 1.01 per mmHg, p <0.001) showed positive associations. However, the protective significant factors includes higher BMI (OR = 0.11 per kg/m^2^, p <0.001), body temperature (OR = 0.57 per °C, p <0.001), and log-transformed white blood cell counts (OR = 0.49, p <0.001) and log-transformed platelet counts (OR = 0.92, p <0.001). Systolic blood pressure (OR = 1.00 per mmHg, p <0.001) and pulse rate (OR = 1.00 per bpm, p <0.001) had negligible effect sizes despite statistical significance. Similarly, the logistic regression analysis of dengue severity indicates that lower platelet counts measured at admissions are linked to a higher likelihood of severe dengue (Table 1 & Fig S1).

**Table 1.** Factors affecting dengue severity. **Left panel:** For clinical predictors with repeated measures during hospital stay, we used a generalized linear mixed-effects model with a binomial link function, accounting for individual patients and different hospitals as random effects. Odds ratios (OR) and p-value are reported. Platelet count and WBC were log-transformed in this model. **Right panel:** For clinical factors measured at the time of admission, we conducted an additional analysis using a logistic regression model. Numerical predictors were normalized to values between 0 and 1, with OR and p-value reported.

|  | Repeated measures during the stay in hospital<br>(Generalized linear mixed effect model) |  | Measures at the time of admission<br>(Logistic regression model) |  |
| --- | --- | --- | --- | --- |
| Factors | OR | p-value | OR | p-value |
| Age | 1.77 | <0.001 | 1.15 | 0.605 |
| BMI | 0.11 | <0.001 | 1.07 | 0.741 |
| Require blood [Yes] | 15.26 | <0.001 | 1.80 | 0.220 |
| SBP | 0.99 | <0.001 | 1.03 | 0.933 |
| DBP | 1.01 | <0.001 | 1.28 | 0.436 |
| Pulse rate | 1.00 | <0.001 | 0.89 | 0.700 |
| Temperature | 0.57 | <0.001 | 1.08 | 0.746 |
| Platelet | 0.92 | <0.001 | 0.46 | 0.033 |
| Hematocrit | 15.16 | <0.001 | 0.74 | 0.189 |
| WBC | 0.49 | <0.001 | 0.94 | 0.798 |

## Discussion

In this observational study, we identified several critical factors associated with severe dengue progression. Older age, blood transfusion requirements, elevated hematocrit levels, reduced platelet counts, and lower body mass index emerged as independent risk factors for disease severity. Clinical timelines, including delays between symptom onset and hospital admission, prolonged hospital stay, and extended time from onset to recovery, were significantly linked to severe outcomes. Additionally, the presence of comorbidities and specific clinical manifestations including headache, abdominal pain, joint pain, respiratory distress, and itchiness were strongly associated with the outcomes of disease severity.

The WHO 2009 guidelines classify dengue progression into three clinical phases—febrile (days 1–3 post-onset), critical (days 3–6), and recovery (>6 days)—with distinct hematological dynamics, including platelet count decline and hematocrit elevation [4]. During the critical phase, platelet counts reach their nadir concurrent with peak hematocrit levels, a hallmark of plasma leakage and thrombocytopenia [14]. Our findings align with these established patterns, showing that log-transformed platelet count reached their lowest point on day 6 post-symptom onset in both groups (Fig 2). Additionally, our results demonstrate that lower platelet counts are strongly associated with progression to severe dengue, reinforcing their role as a key WHO warning sign [4]. This corroborates observational studies in diverse healthcare settings, where thrombocytopenia consistently predicts severe outcomes [15, 16]. In severe dengue cases, the requirement for blood transfusions serves as an important indicator of disease severity and associated complications. Patients often exhibit significant hemorrhagic manifestations due to thrombocytopenia and coagulopathy, leading to an increased need for platelet and red blood cell transfusions [17]. Additionally, plasma leakage can result in hypovolemic shock, necessitating transfusion to restore hemodynamic stability [18]. Studies have shown that higher transfusion requirements correlate with increased morbidity and mortality, underscoring the critical nature of these interventions in managing severe cases [19]. Therefore, monitoring transfusion needs provides valuable insights into disease progression and informs clinical decision-making.

The mean onset-to-admission times and mean recovery times since symptom onset were found to be significantly longer for severe cases (Table S1). The association between clinical timelines and severe dengue outcomes is well-documented in the literature [20-22]. Delays in seeking medical attention after symptom onset can significantly worsen prognosis. A systematic review highlighted that prolonged intervals between symptom onset and hospital admission are linked to increased severity of the disease, as timely intervention is crucial for managing complications such as hemorrhagic manifestations and shock [22]. Moreover, the duration of hospital stay serves as an important indicator of severity. Patients with severe dengue often require longer hospital stays due to the need for intensive monitoring and management of complications [21]. This extended hospital stay not only reflects disease severity but also contributes to increased healthcare costs and resource utilization [21]. Furthermore, the time from symptom onset to recovery is critical; longer recovery times are associated with more severe disease trajectories, as those with significant delays often have underlying complications necessitating prolonged medical care [20]. This emphasizes the need for healthcare systems to enhance awareness of dengue symptoms and improve access to timely medical care to mitigate the risk of severe outcomes.

Our findings indicating an association between lower BMI and severe dengue outcomes contrast with studies reporting obesity as a risk factor for severe manifestations (Table 1 & Table S1). For instance, research in Thailand demonstrated that overweight pediatric patients (BMI ≥25) exhibited a significantly higher prevalence of severe plasma leakage (DHF grades III–IV) compared to those with mild leakage (DHF grades I–II) (45.5% vs. 18.8%) [23]. This discrepancy may reflect population-specific differences in metabolic health, age-related immune responses, or comorbidities. While obesity has been hypothesized to exacerbate dengue severity through mechanisms such as chronic inflammation and endothelial dysfunction [11], our results suggest that undernutrition or metabolic alterations linked to lower BMI could also drive adverse outcomes, potentially via impaired immune competence or delayed recovery. Notably, atypical presentations such as encephalopathy and fluid overload in overweight pediatric cohorts [24, 25] underscore the need for tailored clinical vigilance. However, conflicting evidence from other settings, where no association was observed between obesity and severe dengue or mortality [12], highlights the complexity of BMI as a predictor. These inconsistencies may arise from variations in study design, regional differences in dengue serotypes, or heterogeneous definitions of obesity across populations.

The elevated risk of severe dengue in older adults may reflect a higher prevalence of comorbidities in this population (Table 1). Although our study did not identify statistically significant associations with specific comorbidities—possibly due to limited sample size or heterogeneous profiles—existing literature highlights diabetes mellitus, hypertension, and chronic kidney disease as critical contributors to severe dengue in older individuals [6, 26].

These conditions may exacerbate severity through mechanisms such as impaired immune responses (e.g., immunosenescence) and endothelial dysfunction, which can enhance plasma leakage and thrombocytopenia during the critical phase of infection. For instance, diabetes-associated hyperglycemia may disrupt platelet function, worsening thrombocytopenia, while hypertension could accelerate vascular permeability, explaining the higher proportion of severe dengue cases observed in our findings.

### Limitations

The study used data collected from patients upon hospital admission, providing precise individual level information on laboratory-confirmed dengue cases admitted to four tertiary hospitals in Dhaka, one of the most densely populated city in the world. We identified significant critical factors associated with disease severity outcome for dengue. However, this study has several limitations that should be considered when interpreting the findings. First, the cross-sectional design limits our ability to establish causal relationships between the identified risk factors and severe dengue outcomes. The longitudinal analysis of the data form such study could be interesting to investigate these research questions further. Second, the study was conducted in four tertiary hospitals in Dhaka City during the peak of the 2023 dengue outbreak, which may not free from the biases in generalization of the results to other regions or non-epidemic periods. Third, the sample size, while substantial, may have been insufficient to detect statistically significant associations for certain comorbidities and symptoms, particularly those with low prevalence. Fourth, the reliance on hospital records and patient interviews for data collection may introduce recall bias or incomplete information, especially for variables such as symptom onset and duration. Finally, the classification of BMI and comorbidities was based on self-reported or hospital-recorded data, which may not always be accurate.

## Conclusion

This study highlights critical demographic, clinical, and laboratory factors associated with severe dengue, including older age, blood transfusion requirements, elevated hematocrit, thrombocytopenia, and lower BMI. Delays in hospital admission, prolonged hospitalization, and extended recovery times were also strongly linked to severe outcomes, emphasizing the importance of timely medical intervention. The presence of comorbidities and specific symptoms, such as abdominal pain, respiratory distress, and itchiness, further underscored the complexity of dengue progression. While our findings contrast with some studies regarding the role of obesity in severe dengue, they suggest that undernutrition or metabolic alterations associated with lower BMI may also drive adverse outcomes. These insights underscore the need for tailored clinical management strategies, particularly for high-risk populations such as older adults and those with comorbidities. Additionally, the study highlights the importance of public health initiatives to improve awareness of dengue symptoms and ensure timely access to healthcare, which could mitigate the risk of severe disease. Future research should focus on longitudinal assessments and mechanistic studies to better understand the interplay between metabolic health, immune responses, and dengue pathogenesis, ultimately informing more effective prevention and treatment strategies.

## Data Availability

Data used in this manuscript can be obtained from the corresponding author upon reasonable request.

https://sph.hku.hk/en/Biography/Ali-Sheikh-Taslim

## Acknowledgments

The authors would like to thank Julie Au for her technical assistance. This project was supported by the General Research Funds from the University Grants Committee of Hong Kong (project code 17111124). The funding bodies had no role in the study design, data collection and analysis, manuscript preparation, or the decision to publish. Special thanks to the authorities at Islami Bank Hospital in Kakrail, Mugda, and Mirpur, as well as Enam Medical College in Savar, for granting access to the dengue ward for data collection.

## Author contributions

Conceptualization: MPH, STA

Data curation: MPH, RM, ANK, BA, MAS, SA, MK, TAA, SJT

Formal analysis: MPH, ANK, BA

Investigation: MPH, STA

Resources: RM, FSB, MN, ABS, STA

Visualization: MPH, STA

Writing – original draft: MPH, STA

Writing – review & editing: All authors

## Supplementary

### List of tables

**Table S1.** Associations between demographic characteristics, clinical timelines, and dengue severity in hospitalized patients during the 2023 Dhaka outbreak.

| Characteristic | Severe<br>N = 66<sup>1</sup> | Non-severe<br>N = 383<sup>1</sup> | p-value<sup>2</sup> |
| --- | --- | --- | --- |
| **Age group (years)** |  |  | **0.035** |
| ≤ 18 | 6 (6.4%) | 88 (94%) |  |
| 19-40 | 34 (15%) | 190 (85%) |  |
| 41-60 | 17 (18%) | 76 (82%) |  |
| 60+ | 9 (24%) | 29 (76%) |  |
| Mean (sd) | 39 (18) | 32 (18) | 0.006 |
| **Sex** |  |  | **0.921** |
| Female | 28 (15%) | 160 (85%) |  |
| Male | 38 (15%) | 223 (85%) |  |
| **BMI** |  |  | **0.022** |
| Underweight | 1 (3.2%) | 30 (97%) |  |
| Normal | 21 (13%) | 136 (87%) |  |
| Overweight | 19 (23%) | 64 (77%) |  |
| Mean (sd) | 24.5 (4.1) | 23.4 (4.9) | 0.083 |
| **Occupation** |  |  | **0.063** |
| Business | 12 (23%) | 41 (77%) |  |
| Housewife | 22 (20%) | 90 (80%) |  |
| Job holder | 10 (12%) | 72 (88%) |  |
| Student | 10 (8.5%) | 108 (92%) |  |
| Others | 5 (14%) | 31 (86%) |  |
| **Require blood/platelet transfusion** |  |  | **0.096** |
| Not required | 48 (14%) | 306 (86%) |  |
| Required | 16 (21%) | 60 (79%) |  |
| **Vaccinated for COVID-19** |  |  | **0.076** |
| Unvaccinated | 5 (7.6%) | 61 (92%) |  |
| Vaccinated | 56 (16%) | 294 (84%) |  |
| **Vaccine doses** |  |  | **0.087** |
| none | 5 (7.6%) | 61 (92%) |  |
| single | 3 (25%) | 9 (75%) |  |
| full | 19 (13%) | 132 (87%) |  |
| booster | 34 (18%) | 153 (82%) |  |
| **Onset-to-admission (days)** |  |  | **0.161** |
| ≤ median | 33 (51%) | 155 (41%) |  |
| > median | 32 (49%) | 219 (59%) |  |
| Mean (sd) | 4.71 (1.88) | 3.99 (2.17) | 0.003 |
| **Onset-to-recovery (days)** |  |  | **<0.001** |
| ≤ median | 35 (54%) | 110 (30%) |  |
| > median | 30 (46%) | 256 (70%) |  |
| Mean (sd) | 8.72 (2.31) | 7.68 (2.48) | <0.001 |
| **Duration of hospital stay (days)** |  |  | **0.334** |
| ≤ median | 21 (32%) | 96 (26%) |  |
| > median | 45 (68%) | 272 (74%) |  |
| Mean (sd) | 4.02 (2.26) | 3.61 (1.69) | 0.405 |

<sup>1</sup>n (%); Mean (sd)
<sup>2</sup>Pearson's Chi-squared test; Wilcoxon rank sum test; Fisher's exact test

**Table S2.** Comparing the comorbidities between severe and non-severe dengue cases.

| Characteristic | Severe, N = 66<sup>1</sup> | Non-severe, N = 383<sup>1</sup> | p-value<sup>2</sup> |
| --- | --- | --- | --- |
| COPD or Asthma |  |  | 0.502 |
| Absent | 62 (14%) | 368 (86%) |  |
| Present | 4 (21%) | 15 (79%) |  |
| Hypertension |  |  | 0.232 |
| Absent | 56 (14%) | 344 (86%) |  |
| present | 10 (20%) | 39 (80%) |  |
| Diabetes mellitus |  |  | 0.105 |
| absent | 55 (14%) | 345 (86%) |  |
| present | 11 (22%) | 38 (78%) |  |
| Heart disease |  |  | 0.548 |
| absent | 62 (15%) | 365 (85%) |  |
| present | 4 (18%) | 18 (82%) |  |
| Other |  |  | 0.336 |
| absent | 51 (14%) | 315 (86%) |  |
| present | 15 (18%) | 68 (82%) |  |
| Any comorbidities |  |  | 0.039 |
| absent | 41 (13%) | 285 (87%) |  |
| present | 25 (20%) | 98 (80%) |  |

<sup>1</sup>n (%)<sup>2</sup>Fisher's exact test; Pearson's Chi-squared test

**Table S3.** Comparing the clinical manifestation between severe and non-severe dengue cases.

| Characteristic | Severe, N = 66<sup>1</sup> | Non-severe, N = 383<sup>1</sup> | p-value<sup>2</sup> |
| --- | --- | --- | --- |
| Fever |  |  | 0.471 |
| absent | 1 (25%) | 3 (75%) |  |
| present | 65 (15%) | 380 (85%) |  |
| Body ache |  |  | 0.225 |
| absent | 30 (13%) | 205 (87%) |  |
| present | 36 (17%) | 178 (83%) |  |
| Headache |  |  | 0.046 |
| absent | 29 (12%) | 219 (88%) |  |
| present | 37 (18%) | 164 (82%) |  |
| Anorexia |  |  | 0.262 |
| absent | 38 (13%) | 248 (87%) |  |
| present | 28 (17%) | 135 (83%) |  |
| Abdominal pain |  |  | <0.001 |
| absent | 35 (11%) | 293 (89%) |  |
| present | 31 (26%) | 90 (74%) |  |
| Diarrhoea |  |  | 0.151 |
| absent | 46 (13%) | 298 (87%) |  |
| present | 20 (19%) | 85 (81%) |  |
| Cough |  |  | 0.151 |
| absent | 46 (13%) | 298 (87%) |  |
| present | 20 (19%) | 85 (81%) |  |
| Malaise |  |  | 0.882 |
| absent | 59 (15%) | 340 (85%) |  |
| present | 7 (14%) | 43 (86%) |  |
| Black stool |  |  | 0.318 |
| absent | 62 (14%) | 369 (86%) |  |
| present | 4 (22%) | 14 (78%) |  |
| Rash |  |  | 0.412 |
| absent | 62 (15%) | 348 (85%) |  |
| present | 4 (10%) | 35 (90%) |  |
| Backpain |  |  | 0.372 |
| absent | 52 (14%) | 319 (86%) |  |
| present | 14 (18%) | 64 (82%) |  |
| Joint pain |  |  | 0.026 |
| absent | 43 (13%) | 298 (87%) |  |
| present | 23 (21%) | 85 (79%) |  |
| Dehydration |  |  | 0.492 |
| absent | 58 (14%) | 347 (86%) |  |
| present | 8 (18%) | 36 (82%) |  |
| Respiratory distress |  |  | 0.013 |
| absent | 51 (13%) | 339 (87%) |  |
| present | 15 (25%) | 44 (75%) |  |
| Slurry speech |  |  | 0.471 |
| absent | 63 (15%) | 371 (85%) |  |
| present | 3 (20%) | 12 (80%) |  |
| Itchiness |  |  | <0.001 |
| absent | 53 (13%) | 356 (87%) |  |
| present | 13 (33%) | 27 (68%) |  |
| Psychiatric problem |  |  | 0.599 |
| absent | 66 (15%) | 377 (85%) |  |
| present | 0 (0%) | 6 (100%) |  |
| Gum bleeding |  |  | 0.054 |
| absent | 61 (14%) | 373 (86%) |  |
| present | 5 (33%) | 10 (67%) |  |
| Chest pain |  |  | 0.162 |
| absent | 55 (14%) | 342 (86%) |  |
| present | 11 (21%) | 41 (79%) |  |
| Edema |  |  | 0.471 |
| absent | 65 (15%) | 380 (85%) |  |
| present | 1 (25%) | 3 (75%) |  |
| vomiting |  |  | 0.339 |
| absent | 27 (13%) | 181 (87%) |  |
| present | 39 (16%) | 202 (84%) |  |

<sup>1</sup>n (%)<sup>2</sup>Fisher's exact test; Pearson's Chi-squared test

### Clinical factors measured at admission that affect dengue severity

We constructed a generalized linear model with a binomial link function to assess the impact of clinical and laboratory profiles (measured at admission) on dengue severity. The model was adjusted for demographic characteristics, including age, BMI, and the need for blood or platelet transfusions during the hospital stay.

The logistic regression analysis of dengue severity indicates that only normalized platelet counts are significantly associated with disease severity (Fig S1). Specifically, for each 1-unit decrease in normalized platelet levels, the odds of a patient experiencing severe dengue (as opposed to non-severe dengue) increase by approximately 54% (as indicated by the odds ratio of 0.46). This suggests that lower platelet counts are linked to a higher likelihood of severe dengue. No other clinical factors were found to significantly affect dengue severity.

**Figure S1.**
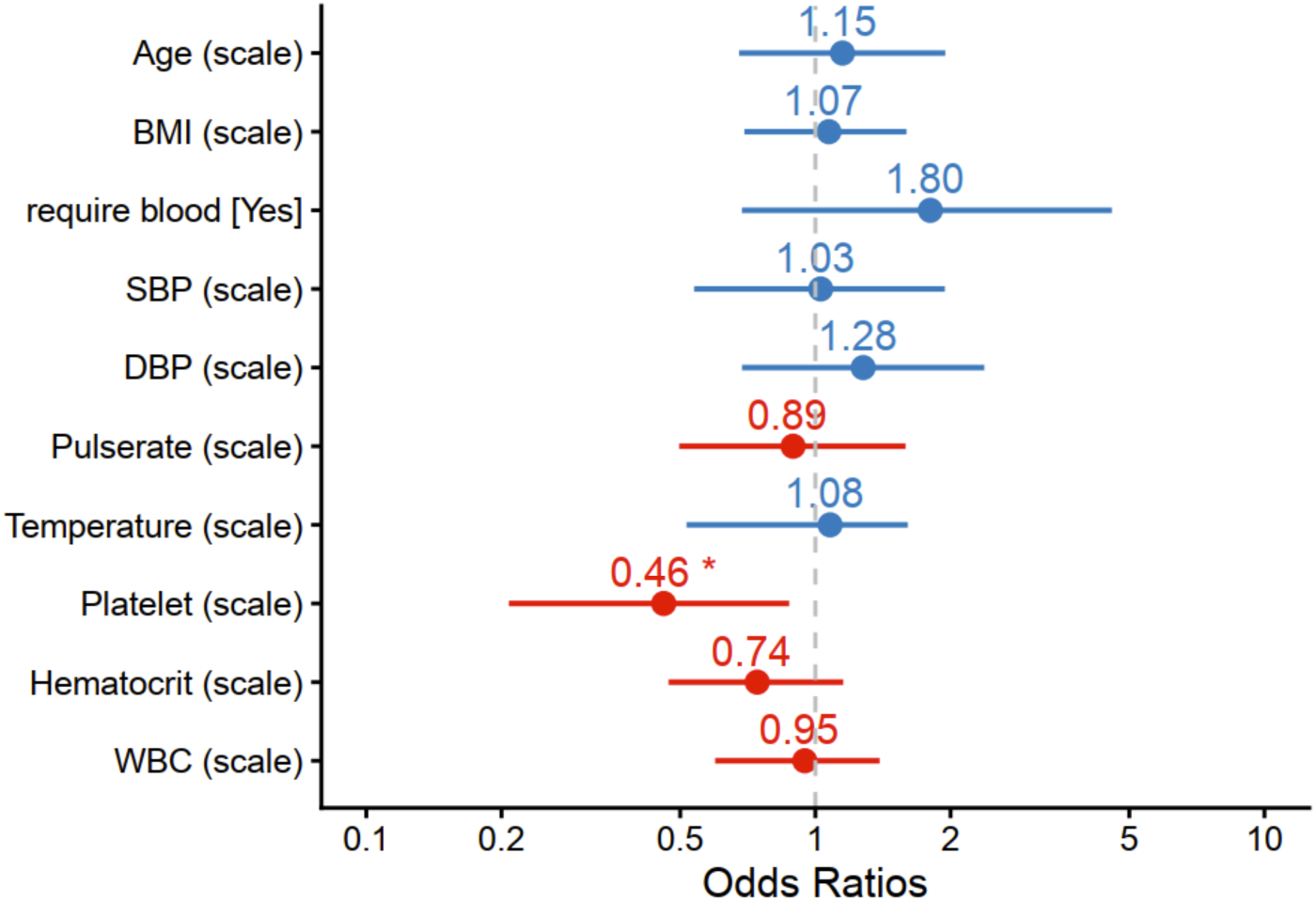
Logistic regression model output illustrating potential clinical factors measured at the time of admission that affect dengue severity. The X-axis represents the odds ratios, while the Y-axis displays the predictors. Numerical predictors were normalized to address variability in their original values, indicated by “(scale)” in parentheses. The red color of the platelet variable indicates a negative association with severe dengue, whereas the blue variables indicate positive associations. Values marked with asterisks denote statistically significant associations.

